# Prevalence and factors associated with short birth interval in Kaya Municipality, North Central Burkina Faso: a multilevel Poisson regression modeling with a robust variance of a community survey

**DOI:** 10.1101/2024.03.18.24304505

**Authors:** Abou Coulibaly, Adama Baguiya, Bertrand Ivlabèhiré Meda, Tiéba Millogo, Aristide Marie Arsène Koumbem, Franck Garanet, Seni Kouanda

## Abstract

A short birth interval adversely affects the health of mothers and children. This study aimed to measure the prevalence of short birth intervals and identify their associated factors in a semi-urban setting in Burkina Faso. We conducted a cross-sectional study in which data were collected in households between May and October 2022. The dependent variable was the short birth interval (SBI), defined by the World Health Organization as the time between two live births. We performed a multilevel mixed-effects Poisson regression with robust variance to determine the factors associated with the SBI by reporting adjusted prevalence ratios (aPR) with a 95% confidence interval (CI). A total of 5544 birth intervals were recorded from 4067 women. A short birth interval was found in 1503 cases out of 5544, i.e., a frequency of 27.1%. The prevalence of short birth interval (time between two live births less than 33 months) was higher in never users of modern contraceptive users (aPR=1.24; 95% CI [1.14-1.34] vs. previous users), in younger ages with aPR of 4.21 (95% CI [3.30-5.37]), 2.47 (95% CI [1.96-3.11]), and 1.45 (95% CI [1.16-1.81]), respectively for under 18, 18-24 years old, and 25-34 years old, compared to 35 and over. Childbirths occurring before the implementation of the maternal and infant free health care policy (aPR=2.13; 95% CI [1.98-2.30]) and also before the free FP policy (aPR=1.53; 95% CI [1.28-1.81]) were found also protective against SBI. Women with low socio-economic positions were more likely to have SBI. This study found a high SBI in Burkina Faso (more than one woman out of four). Our results have programmatic implications, as some factors, such as contraceptive practice and socioeconomic status, are modifiable. These factors need particular attention to lengthen birth intervals and, in turn, improve mother-child couple health by reducing short birth interval consequences.

## Introduction

The short birth interval (time between two live births less than 33 months) has health consequences for both mother and child. It has been shown that a short birth interval, compared with a longer one (greater than or equal to 33 months), increases neonatal, infant, and child mortality by 85%, 116%, and 126%, respectively [1]. Despite the implementation of several strategies to improve maternal and infant health in many countries around the world [2–4], mortality rates remain very high globally. According to the United Nations Inter-agency Group for Child Mortality Estimation, if current trends continue, 48·1 million under-5 deaths are projected to occur between 2020 and 2030, almost half of them projected to occur during the neonatal period [5]. One of the factors contributing to these high mortality rates is the short birth interval, which could result in eclampsia, uterine rupture, post-partum hemorrhage, severe anemia, or even maternal death [6]. The hypertensive and hemorrhagic consequences may account for more than 50% of maternal mortality worldwide [7].

According to the literature, there are several factors associated with short birth interval: the duration of breastfeeding of the previous child, the number of antenatal care (ANC) visits for the index pregnancy, woman’s level of education, woman’s occupation, contraceptive practice, woman’s age at marriage, the woman’s age at first pregnancy, partner’s religion, partner’s level of education, partner’s occupation, the parity of the index child, the survival of the previous birth, the place of residence and the well-being index, etc. [8–11]. Most studies have considered only the woman’s most recent birth interval and therefore excluded her previous birth intervals. Some other studies did not use the World Health Organization (WHO) definition of a short birth interval, thus limiting the comparability of the findings across studies, as Islam et al. showed in their published systematic review and meta-analysis [12]. Also, a multicenter study in eight African countries concluded that the determinants of the short birth interval varied from one country to another [13]. Pimentel et al., in a systematic review, noted that shorter breastfeeding and the female sex of the previous child were the only factors systematically associated with a short birth interval. They concluded that future quantitative research should use longitudinal and experimental designs and ensure consistency of outcome and exposure definitions in the search for determinants of short birth intervals [14].

Considering the limitations of previous studies, this study aimed to estimate the prevalence and identify the factors associated with the short birth interval in a semi-urban Municipality in Burkina Faso using the WHO definition and including all women’s birth intervals.

## Materials and Methods

### Study setting

The study was conducted in the Kaya Demographic and Health Surveillance System (Kaya-HDSS) catchment area in Kaya municipality, North Central, Burkina Faso. This demographic and health surveillance site has been described elsewhere [15]. It has been operational since 2007 and is managed by the Institut de Recherche en Sciences de la Santé (IRSS). According to the latest General Census of Population and Housing, in 2019, Kaya’s population was estimated at 208,682 [16]. In 2022, the results of the Kaya SSDS showed a resident population of 82,723, including 21,854 women aged 15 to 49 and 4,888 children under 5. This population-based observatory covers one medical center, six urban primary health centers known as the Centre de Santé et de Promotion Sociale (CSPS), six rural health centers, and some private health centers. In addition to these primary health centers, the observatory area is also home to the Regional Hospital (CHR), which provides specialist care and acts as a referral center for the health centers in the four provinces of the Centre-Nord region. The Kaya demographic and health surveillance site aims to study demographic, infectious, and chronic disease indicators in the district, observe changes in health over time, evaluate health programs, and provide a basis for policy decisions and capacity to improve the health of the community [17].

### Study design and period

We conducted a population-based cross-sectional study. Data were collected between May and October 2022 in the demographic and health surveillance system catchment area.

### Study population

The study population consisted of women of childbearing age (15 to 49 years) living in the Kaya demographic and health surveillance site.

### Inclusion criteria

This study included women who had had at least two consecutive live births in the previous ten years (2012-2022) and were present in the households when the interviewers visited.

### Study variables

The dependent variable in our study was the birth interval for two consecutive live births, a binary variable coded ’1’ for a birth interval of less than 33 months (considered as short) and ’0’ for a birth interval of 33 months or more (considered as optimal), based on the WHO definition [18]. We used all children’s dates per woman from 2012 to 2022. Due to the hierarchical type of the variables, we considered trois levels: (i) the delivery level, the woman level (woman could have many deliveries), and the village level. For each level, independent variables were defined, as shown in Table 1.

**Table 1:** level-related independent variables.

| Level | Independent variables | Modalities |
| --- | --- | --- |
| level 1: deliveries | Childbirth during free maternal and child health care policy implementation | (Yes or No) |
|  | Childbirth before free family planning policy implementation | (Yes or No) |
|  | Pregnancy rank | (from 2 to 10) |
|  | Women's age at the start of pregnancy (in years) | Under 18, 18-24, 25-34, 35 and over |
| Level 2: woman (variable woman id) | Woman marital status (in union) | (Yes or No) |
|  | Wealth quintiles | Lowest, Second, Middle, Fourth, Highest |
|  | Ever used a modern method of contraception | (Yes or No) |
| Level 3: village level | Place of residence | Urban, Rural |

### Data source and collection

Data were collected through individual interviews with women who met the inclusion criteria and were present in the households when the interviewers visited. Some data were extracted from women’s health records and children’s birth certificates. The questionnaire included variables such as past and current contraceptive use, the outcome of each pregnancy, the survival of each child, and so on. The data from this study were supplemented by data from the Kaya-HDSS, which are repeated cross-sectional study data. The data extracted from Kaya-HDSS mainly concerned the socio-demographic characteristics of women and their households, socio-economic level, etc.

### Data analysis

The data were analyzed using Stata software version 18.0 [19]. Qualitative variables were described using frequencies. Because of the data structure (women have multiple birth intervals), we performed a multilevel mixed-effects Poisson regression with robust variance to determine the factors associated with the short birth interval by reporting prevalence ratios (PR) with a 95% confidence interval rather than odds ratios to avoid a poor approximation of risk because the prevalence found was not low [20–22]. After fitting the null model (model 1), we first included the level-1 variables (model 2), then the level-2 variables (model 3), and lastly, the level-3 variables (model 4). These models are available in Supplementary File 1. The results of the final multivariable model are shown in Table IV, and its selection was based on the Akaike Information Criteria (AIC) and Bayesian Information Criteria (BIC). All statistical tests were two-sided; a p-value < 0.05 was considered statistically significant.

### Ethical considerations

The study protocol was approved by the Burkina Faso Health Research Ethics Committee (Deliberation Number 2021-07-165). Informed consent was obtained from each participant before the questionnaire was administered. Women’s anonymity and confidentiality were ensured throughout data collection and processing.

## Results

Four thousand one hundred eight women had at least two consecutive live births between 2012 and 2022 and were included in this study. Of these, 40 were excluded because of inconsistent data on birth dates. In total, 5,544 birth intervals were analyzed. Fig 1 below shows the flow chart.

**Figure 1:**
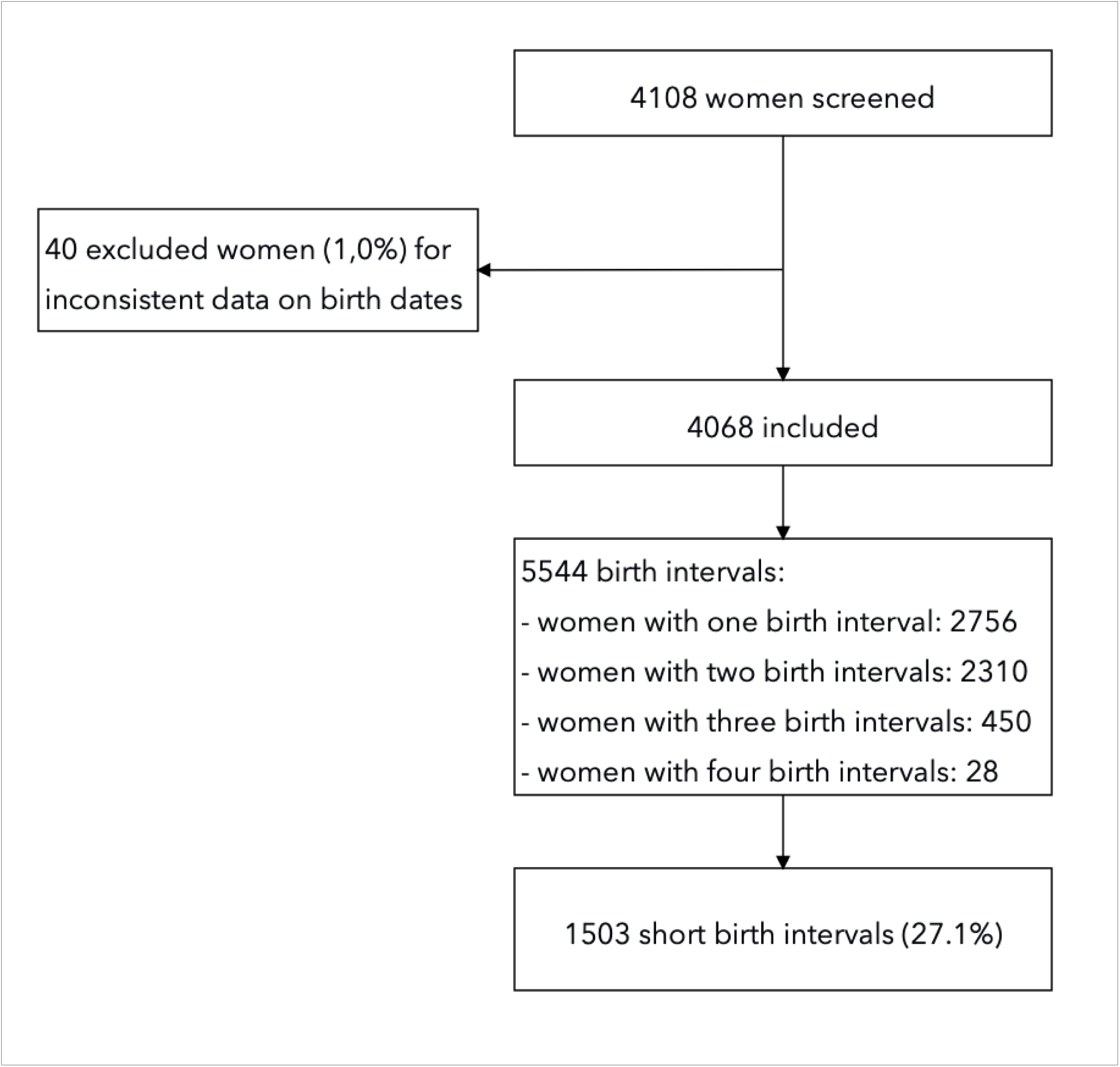
study flow chart.

### Characteristics of pregnancies and deliveries

Out of the deliveries, 81.5% occurred during maternal and infant-free healthcare (after 1 June 2016), and 13.3% of pregnancies began during free family planning (after 1 July 2020), as detailed in Table 2.

**Table 2:** Characteristics of pregnancies and births (N=5544)

| **Characteristics of pregnancies and births** | **N=5544** | **%** |
| --- | --- | --- |
| **Age at the start of pregnancy** |  |  |
| Under 18 | 326 | 5,9 |
| 18-24 | 2233 | 40,3 |
| 25-34 | 2472 | 44,6 |
| 35 and over | 513 | 9,3 |
| **Pregnancy rank (first to last pregnancy)** |  |  |
| 2 | 1521 | 27,4 |
| 3 | 1329 | 24,0 |
| 4 | 1062 | 19,2 |
| 5 | 758 | 13,7 |
| 6 | 459 | 8,3 |
| 7 | 222 | 4,0 |
| 8 | 109 | 2,0 |
| 9 | 58 | 1,0 |
| 10 and over | 26 | 0,5 |
| **Childbirth during maternal and child free healthcare policy** |  |  |
| No | 1025 | 18,5 |
| Yes | 4519 | 81,5 |
| **Pregnancy during free family planning policy implementation** |  |  |
| No | 4809 | 86,7 |
| Yes | 735 | 13,3 |

### Characteristics of the women

Women who had ever used a modern method of contraception were 71.0%, and 95.7% of women were in union. Table 3 shows the socio-demographic characteristics of the women.

**Table 3:** Socio-demographic characteristics of the women.

| **Socio-demographic characteristics** | **N=4068** | **%** |
| --- | --- | --- |
| **Main occupation** |  |  |
| Housewife | 2235 | 54,9 |
| Farmer | 749 | 18,4 |
| Shopkeeper | 964 | 23,7 |
| Student/Public or private worker | 120 | 2,9 |
| **Woman in union** |  |  |
| No | 174 | 4,3 |
| Yes | 3894 | 95,7 |
| **Place of residence** |  |  |
| Urban | 2479 | 60,9 |
| Rural | 1589 | 39,1 |
| **Wealth quintiles** |  |  |
| Lowest | 955 | 23,5 |
| Second | 806 | 19,8 |
| Middle | 783 | 19,2 |
| Fourth | 776 | 19,1 |
| Highest | 748 | 18,4 |
| **Ever used a modern method of contraception** |  |  |
| No | 1178 | 29,0 |
| Yes | 2890 | 71,0 |

### Prevalence of short birth interval and associated factors

A short birth interval was observed in 1503 cases out of 5544, i.e., a frequency of 27.1% (95% CI: 26.0-28.3). Table 4 shows the factors associated with a short birth interval in univariable and multivariable analyses. For example, the adjusted analyses showed that the prevalence of short birth interval was increased by 24% in women who had never used a modern contraceptive method (aPR=1.24; 95% CI [1.14-1.34]) compared with those who ever used it. Women aged under 18 during their pregnancies, and those aged 18-24, and 25-34, were respectively, 4.21, 2.47, and 1.45 times more likely to experience a SBI, compared with those aged over 35. Other factors, such as pregnancy rank, wealth quintiles, the infant and mother free healthcare policy, and the free family planning policy, were strongly associated with the short birth interval.

**Table 4:** Results of univariate and multivariate analyses (N=5544)

|  | **Crude**<br>**Prevalence**<br>**Ratio (95% CI)** | **p** | **Adjusted**<br>**Prevalence**<br>**Ratio (95%**<br>**CI)** | **p** |
| --- | --- | --- | --- | --- |
| **Childbirth during women and**<br>**infant and mother free healthcare**<br>**policy** |  |  |  |  |
| No | 2.53(2.34-2.75) | <0.001 | 2.13(1.98-2.30) | <0.001 |
| Yes |  |  |  |  |
| **Pregnancy during free family**<br>**planning policy implementation** |  |  |  |  |
| No | 2.04(1.71-2.42) | <0.001 | 1.53(1.28-1.81) | <0.001 |
| Yes |  |  |  |  |
| **Pregnancy rank** |  |  |  |  |
| 2 | 0.76(0.42-1.35) | 0.347 | 0.45(0.24-0.85) | 0.013 |
| 3 | 0.66(0.38-1.14) | 0.138 | 0.49(0.27-0.88) | 0.018 |
| 4 | 0.61(0.33-1.12) | 0.109 | 0.55(0.29-1.05) | 0.069 |
| 5 | 0.56(0.32-0.98) | 0.044 | 0.59(0.33-1.05) | 0.074 |
| 6 | 0.60(0.32-1.11) | 0.106 | 0.67(0.36-1.25) | 0.210 |
| 7 | 0.72(0.41-1.25) | 0.237 | 0.77(0.46-1.29) | 0.324 |
| 8 | 0.83(0.47-1.47) | 0.521 | 0.99(0.55-1.78) | 0.975 |
| 9 | 1.06(0.61-1.83) | 0.845 | 0.93(0.56-1.54) | 0.766 |
| 10 and over |  |  |  |  |
| **Age at pregnancy** |  |  |  |  |
| Under 18 | 3.06(2.50-3.76) | <0.001 | 4.21(3.30-5.37) | <0.001 |
| 18-24 | 1.63(1.34-1.98) | <0.001 | 2.47(1.96-3.11) | <0.001 |
| 25-34 | 1.13(0.92-1.39) | 0.257 | 1.45(1.16-1.81) | 0.001 |
| 35 and over |  |  |  |  |
| **Woman in union** |  |  |  |  |
| No |  |  |  |  |
| Yes | 0.73(0.60-0.87) | 0.001 | 0.85(0.71-1.02) | 0.077 |
| **Wealth quintiles** |  |  |  |  |
| Lowest | 1.49(1.26-1.76) | <0.001 | 1.30(1.04-1.62) | 0.020 |
| Second | 1.50(1.36-1.65) | <0.001 | 1.28(1.12-1.47) | <0.001 |
| Middle | 1.35(1.19-1.54) | <0.001 | 1.18(1.01-1.38) | 0.040 |
| Fourth | 1.08(0.94-1.24) | 0.289 | 1.02(0.87-1.19) | 0.850 |
| Highest |  |  |  |  |
| **Ever used a modern method of contraception** |  |  |  |  |
| No | 1.32(1.22-1.44) | <0.001 | 1.24(1.14-1.34) | <0.001 |
| Yes |  |  |  |  |
| **Place of residence** |  |  |  |  |
| Urban |  |  |  |  |
| Rural | 1.16(1.01-1.34) | 0.037 | 0.91(0.79-1.03) | 0.143 |
| **Main occupation** |  |  |  |  |
| Housewife | 1.42(1.15-1.74) | 0.001 |  |  |
| Farmer | 1.42(1.07-1.90) | 0.016 |  |  |
| Shopkeeper | 1.04(0.80-1.35) | 0.764 |  |  |
| Student/Public or private worker |  |  |  |  |

## Discussion

The proportion of short birth intervals was 27.1% in our study. This is high in a context where many actions are being undertaken to promote modern contraceptive methods. Some of the factors found in our study can be addressed by implementing or reinforcing specific health policies. For example, our results show that the prevalence of the short birth interval has significantly decreased after the start of free family planning policy nationwide in 2019. This policy, therefore, needs to be monitored to ensure that it continues to be effective with women. This high prevalence should raise questions and lead to a strengthening of health policies aimed at birth spacing.

This high prevalence is common in many African countries. It has also been noted in the literature, ranging from 16.99% to 58.74%: analyses carried out on Demographic and Health Survey (DHS) databases showed that the pooled prevalence was 16.99% for 12 East African countries in a study published in 2023 [10], and 58.74% as a pooled prevalence in 10 African countries with high fertility [23]. Another study in 35 sub-Saharan African countries found a pooled prevalence of 43.91%, with South Africa having the lowest prevalence (23.25%) and Chad having the highest (59.28%). In the same study, the authors also noted that 14 of the 35 countries had a higher prevalence than the sub-Saharan African average of 43.91% [24]. Several community-based cross-sectional studies carried out in Ethiopia give similar high prevalence, such as 56% in 2020 [25] and 30.59% in the north-west in 2021 [26] Our study has shown that women’s age at the pregnancy, contraceptive use in the woman’s reproductive life, socioeconomic status of the woman, and pregnancy rank were associated with SBI. Several studies [27–29] have also found similar results. Therefore, women who practice family planning and can intentionally choose to space their births are logically less likely to experience a short birth interval.

Our results also showed that the wealth index was associated with a short birth interval: the prevalence of a short birth interval was higher among women in the poor and very poor quintiles than their counterparts in the rich and very rich quintiles. This result supports the conclusions of several other studies [30–33]. Women in the wealthiest groups had greater access to quality primary care than women in the poorest group. For example, women in the rich and very rich quintiles would benefit from more prenatal contacts. However, an increase in antenatal visits has been shown to reduce the risk of having a short birth interval [34]. Furthermore, women in the rich and very rich quintiles may be better educated than those in the poorer groups. Given the strong link between education level and desire for children (negative association between formal schooling and desired fertility) [35,36], they could, therefore, space births better than poor women. However, a study in Ethiopia showed that poor households were less likely than wealthy households to have a short birth interval [37]. This author nevertheless pointed out the presence of recall bias in his study concerning the actual interval between births and bias in the measurement of the wealth index, which could explain his results, which were at odds with ours and the data in the literature.

In our study, the place of residence did not determine the birth interval length. While some studies have shown no difference between rural and urban women regarding the length of the birth interval [38,39], others have shown that rural women are more likely to have a short birth interval than urban women [32,40,41]. A difference in the composition of the place of residence in these countries could justify the differences between these results. Furthermore, in our study, the Nord Central region, where the Kaya-HDSS is located, has been affected by security issues for nearly eight years now, and many rural women from the rural areas have moved to the city of Kaya. Therefore, the place of residence variable is subject to significant misclassification, which may explain the lack of association between the place of residence and the short birth interval.

Finally, it should be noted that the short birth interval may be the result of a deliberate choice on the part of the woman or even the couple: in Nigeria, in 2010, a study reported as its main findings the fact that the women attributed responsibility for a high parity to their husbands. They had deliberately given birth to many children to inhibit men’s tendency to divorce or enter into multiple marriages [42].

The main strengths of this study are (i) its design, which uses data from all women’s birth intervals over ten years, and (ii) the multilevel mixed-effects Poisson regression with robust variance we performed to determine the factors associated with the short birth interval by reporting prevalence ratios, compared to previous studies. The main limitation is the recall bias that arises when women are asked about past events, such as past childbirths, if the existing documentation does not allow the event to be located accurately.

## Conclusion

At the end of this study, we found that the prevalence of short birth intervals is still high in Burkina Faso (more than one woman in four). Our results have programmatic implications because some factors, such as contraceptive practice and socioeconomic status, are modifiable. These factors merit particular attention to lengthening birth intervals and, in turn, reduce their consequences by improving the health of the mother-child couple. Activities to encourage contraceptive use should, therefore, be stepped up to increase the number of new users and retain existing users. These users will increase their chances of having an appropriate birth interval.

## Authors’ contributions

Conceptualization: AC. Methodology: AC. Supervision of data collection: AC, AB, FG. Validation: AC, AB, FG. Formal analysis: AC, AB, BIM, TM. Writing-original draft preparation: AC. Writing-review and editing: AC, AB, BIM, TM, AMAK. Project administration: AB, SK. Funding acquisition: SK. All authors have read and agreed to the published version of the manuscript.

## Data Availability

The datasets used and analyzed during the current study are available from the corresponding author upon reasonable request.

## Acknowledgements

We sincerely thank Fatou Sissoko, Vincent Bagnoa, Bouraima Kabré, Ambroise Bamogo and all the interviewers for their support during data collection.

